# Molecular identification of species of family Chaetomiaceae (Sordariomycetes, Ascomycota) from soil, dung and water in Sudan

**DOI:** 10.1101/2022.04.05.22273477

**Authors:** Najwa A Mhmoud

**Affiliations:** Faculty of Medical Laboratory Sciences, Department of Medical Microbiology and Immunology. University of Khartoum. Khartoum. Sudan

**Keywords:** Chaetomiaceae, Indoor species, Morphological diversity, Phylogeny

## Abstract

Species of Chaetomiaceae family are ubiquitous filamentous fungi, that responsible for a wide range of opportunistic human infections. To date, it encompasses more than 300 species have been described in genus Chaetomium. It have been globally reported as being are capable of colonizing various substrates. Due to the lack of genetic studies on the species belonging to this genus in Sudan, This work aimed to investigate the environmental fungal occurrence within the family Chaetomiaceae by using morphological characters and molecular sequencing.

A total of 260 environmental samples from soil, animal dung and water were collected from six different states in Sudan in two ecozones: .desert or semis desert ecozones (Dongola in Northern sudan, El-Obeid in western sudan); a low rainfall woodland savanna ecozone (Gazira, El Geteina and Khartoum from central Sudan and AlQadarif in eastern Sudan).

During a study of environmental fungi in Sudan, 119 isolates were identified as members of Chaetomiaceae after the ITS sequencing combined with an examination of the macro- and micromorphology. Out of 63 Chaetomium strains obtained from soil, animal dung and water samples, 25 were obtained from soil, 22 from animal dung and 16 from water. 56 additional strains isolated from other genus within the Chaetomiaceae family, such as (*Amesia, Collariella, Ovatospora, Subramaniula* and *Thielavia*) were recorded for the first time in Sudan.

In conclusion: Sequence-based identification of fungal isolates is often considered to be the most reliable and accurate identification method.

## Introduction

The genus, Chaetomium, which belongs to *Chaetomiaceae* family and Sordariales order (class Sordariomycetes in Ascomycota) was first recognized and established in 1817, with *Chaetomium globosum*, the type species of the genus, was first described by Kunze [1]. Then after that the taxonomy of Chaetomium has been studied by several authors [2–11]. To date, it encompasses more than 300 species have been described in genus Chaetomium. It have been globally reported as being are capable of colonizing various substrates and are well-known for their ability to degrade cellulose and to produce a variety of bioactive metabolites [12–15]. As cellulose-degrading fungi they possess the ability to degrade cellulosic building materials such as wood, plastics and drywall [16]. These had adverse effects not only on the buildings but also to their occupants [17]. It can cause minor disorders such as allergic sinusitis, asthma [18]. onychomycosis [19–26], superficial skin mycosis [24], cerebral mycosis [27–29] and recently can cause eumycetoma [30].

The genus Chaetomium is generally characterized by superficial rounded, ovoid, ostiolate ascomata covered with characteristic hairs. Ascomatal hairs can be straight (seta-like), flexible, curved, wavy, circulating, spirally curled, or otherwise branched in various morphologies. Asci are clavate or fusiform with 8 biseriate or irregularly arranged ascospores. Ascospores are limoniform to globose, or irregular in a few species, bilaterally flattened, brown or gray –brown (never opaque or black) [10].

The first well classification system of Chaetomium based on both the morphological characters (ascogonia, asci, ascospores and anamorphs) and physiological requirements (temperature, nutritional requirements, growth and fruiting rate) was established by Dreyfuss In 1976 [31] Among several attempts to accommodate Chaetomium in a higher-rank taxonomy, Winter (1885) was the first to include the genus in family Chaetomiaceae (Sordariales, Sordariomycetes), a placement that has hold out all critical tests [32]. Then great effort has been made to classify, identify, and accurately grouping different species of *Chaetomium*, based on DNA sequencing [18]. Chaetomium and other members of the Chaetomiaceae were studied with 18S and 28S rDNA sequences individually [33, 34] (or in combination with tef and rbp2 genes [35] These studies have supported the monophyly of the Chaetomiaceae, occupying a sister relationship to other families of Sordariales, especially Lasiosphaeriacea. Some *Chaetomium* species are reported to be associated with *Acremonium, Botryotrichum, Humicola, Paecilomyces*, and *Scopulariopsis* [36]. In addition to that the distinctions among Achaetomium, chaetomidium, chaetomium and thielavia were un clear [37]. Therefore additional molecular studies on this genus are necessary [18]. Due to the lack of genetic studies on the species belonging to this genus in Sudan, This work aimed to investigate the environmental fungal occurrence within the family Chaetomiaceae by using morphological characters and molecular sequencing.

## MATERIALS AND METHODS

### Sample collection

A total of 260 environmental samples from soil, animal dung and water were collected from six different states in Sudan from two ecozones: .desert or semis desert ecozones (Dongola in Northern sudan, El-Obeid in western sudan); and a low rainfall woodland savanna ecozone (Gazira, El Geteina and Khartoum from central Sudan and AlQadarif in eastern Sudan) (figure-1).

**Figure 1:**
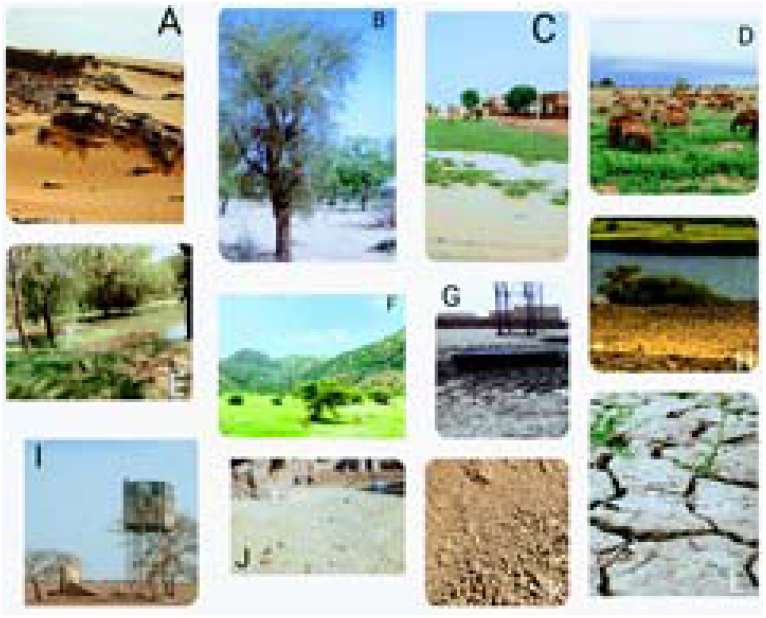
Sampling location in Sudan States : A. Dongola in Northern Sudan cover by Yermosol (Desert sand); B El-Obeid in Sahelian Acacia savanna zone with Arenosols (Stabilized sand dunes with silt or clay soil); C. Gezira state in Sahelian Acacia savanna zone with Vertisols (Black clay soil); D. El Geteina city in Sahelian Acacia savanna zone with Yermosols (Flat gravel silt loam with clay soil ; E. Khartoum city in Sahelian Acacia savanna zone with Yermosols (Flat gravel silt loam with clay soil); F. AlQadarif in East Sudanian savanna with Vertisols (Black clay) ; G, Animal drinking water; H. Surface river water sample; I. Drinking water sample; J. Animal manure K. loamy soil sample; L. Clay soil sample.

Dongola is the capital of the state of Northern Sudan, on the banks of the Nile. It has an area of 348,765 km^2^.(figure-1A).. El-Obeid is the capital of the North Kordofan states. It has an area of 130 km^2^ (figure-1B). Gezira state lies between the Blue Nile and the White Nile in the east-central region of the country. It has an area of 23.373 km^2^ (figure-1C). El Geteina is a small town located in the state of the White Nile Sudan. It has an area of 6602 km^2^ (figure-1D). Khartoum city is located in the heart of Sudan at the confluence of the White Nile and the Blue Nile, where the two rivers unite to form the River Nile. It has an area of 28165 km^2^ (figure-1E). AlQadarif is the capital of the state. It has an area of 75,263 km^2^ (Figure-1F).

### Soil samples collection

Approximately 150-200 g was collected from each soil sample at a depth of 0-10 cm after scraping and removing leaves and other plant debris on the soil surface (Figure-1k and L). A total of one hundred and twenty Soil samples were collected and were put in clean bags.

### Animal dung collection

A total of ninety dung samples were collected from horse, donkey, sheep and cow manure(Figure-1J). The samples are placed in sterile sealed containers.

### Water samples collection

Fifty water samples have been randomly collected in sterile bottles (500ml) from surface river water (Figure-1H), drinking water (Figure-1I), sewage water and animal drinking water(Figure-1G)..

All the samples were then transferred to the laboratory, and maintained at 4°C until further use.

### Measurements of Physicochemical Parameters

We employed the Sudan Soil Information System (SUSIS) digital soil map of the Sudan from Land and Water Research Centre - Agricultural Research Corporation Sudan as a reference on the type of soil where the species occurred. Soil important characteristics, Soil texture, salinity, pH and nitrogen, carbon, calcium carbonate potassium and phosphorus concentrations were retrieved from SUSIS database (Figure-2, 3 and 4).

**Figure 2:**
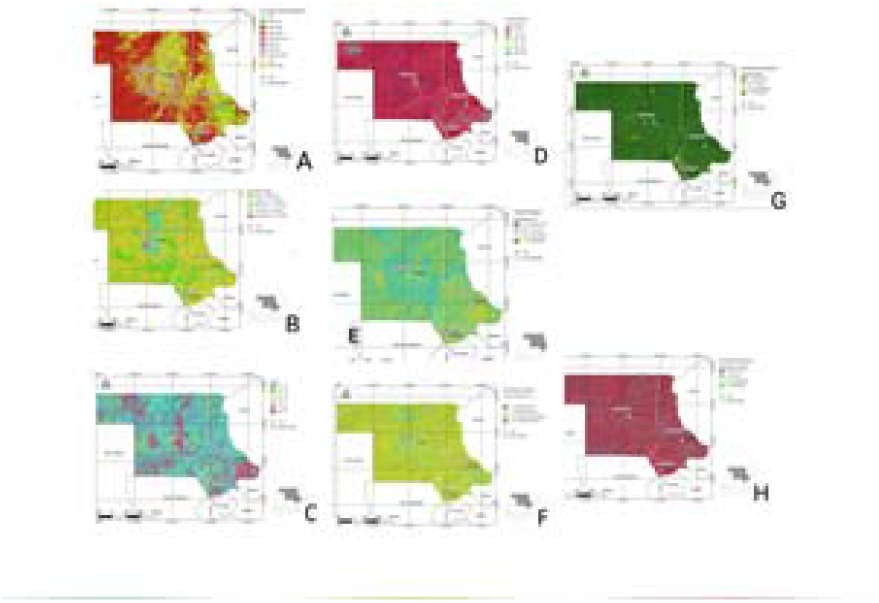
Soil characteristic of Dongola and Khartoum state : A. Soil texture; B. Soil salinity; C. Soil pH.; D. Carbon concentrations, ; E. Nitrogen concentrations F. Calcium carbonate concentrations, G. Soil potassium ; H. phosphorus concentrations. Figure was created for this publication using from Sudan Soil Information System (SUSIS) digital soil map of the Sudan from Land and Water Research Centre - Agricultural Research Corporation Sudan

**Figure 3:**
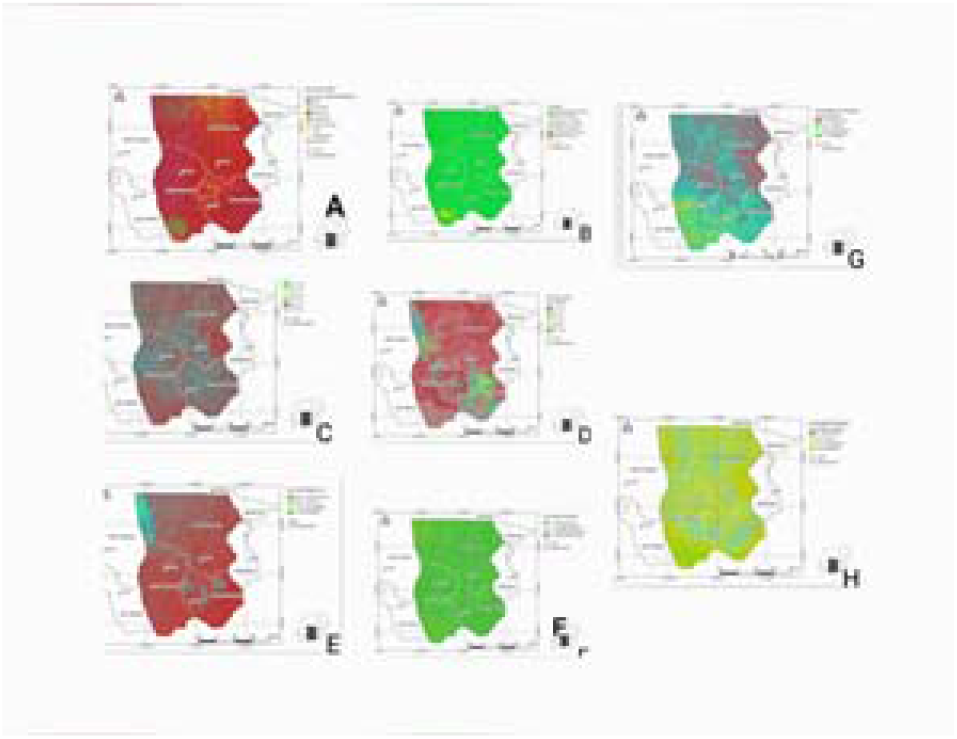
Soil characteristic of El-Obeid the capital of the North Kordofan states : A. Soil texture; B. Soil salinity; C. Soil pH.; D. Carbon concentrations, ; E. Nitrogen concentrations F. Calcium carbonate concentrations, G. Soil potassium ; H. phosphorus concentrations. Figure was created for this publication using from Sudan Soil Information System (SUSIS) digital soil map of the Sudan from Land and Water Research Centre - Agricultural Research Corporation Sudan

**Figure 4:**
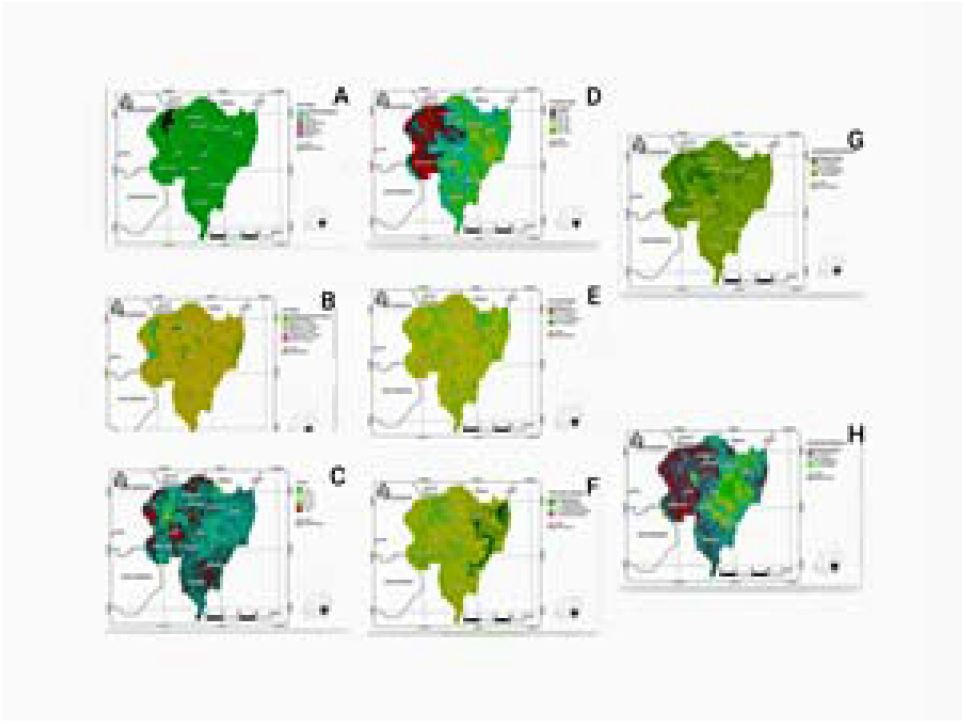
Soil characteristic of AlQadarif region El Geteina city and Gezira states: A. Soil texture; B. Soil salinity; C. Soil pH.; D. Carbon concentrations, ; E. Nitrogen concentrations F. Calcium carbonate concentrations, G. Soil potassium ; H. phosphorus concentrations. Figure was created for this publication using from Sudan Soil Information System (SUSIS) digital soil map of the Sudan from Land and Water Research Centre - Agricultural Research Corporation Sudan

### Isolation of fungi

The fungi within the family Chaetomiaceae were isolated from soil using the plate dilution method [38]. Briefly, from each soil sample a total of 10 g was suspended in 100ml sterile distilled water and shaking for 10 min at 25C° and 120 rpm. The suspensions of soil were serial diluted in sterile distilled water up to 10^−6.^approximately 1ml was removed by pipette and streaked.

For primary isolation, potato dextrose agar (PDA; Hi media, India), Sabouraud dextrose agar (SDA; Hi media, India) and oatmeal agar (OA; Hi media, India) supplemented with the antibiotic chloramphenicol (50 mg/L) were used with three replicates. All the dishes were incubated at 25 °C, 37 °C and 45 °C in the dark for 7-14 days. All distinct colonies were subjected to additional purification by sub culturing onto PDA and Malt extract agar, (MEA 2%; Merck, Germany), and the plates were incubated for 5–7 days at 25 °C, 37 °C and 45 °C in the dark until growth of fungal colonies was observed. Morphologically distinct colonies were selected and further purified by sub-culturing on PDA and MEA. The pure cultures were preserved on 20% glycerol stock at 4°C for further studies.

For animals dung, the alcohol treatment technique was used to isolate fungi from the dung samples. This technique was undertaken to stimulate ascospore germination on agar media [39]. Each 1 g sample was soaked in 9 ml of 65% ethyl alcohol in a test tube and incubated at 25C° and 30°C for 15 min. Samples were mixed thoroughly and then spread onto PDA, SDA and OA supplemented with the antibiotic chloramphenicol (50 mg/L) were used with three replicates. All the dishes were incubated at 25 °C, 37 °C and 45 °C in the dark for 7-14 days.

Furthermore strains were isolated from water samples using filtration technique [40]. One hundred milliliters of each water sample was filtered through membrane filters (pore diameter 0.45 μm), followed by incubation of the membrane filters on PDA, SDA and OA supplemented with the antibiotic chloramphenicol (50 mg/L) were used with three replicates. All the dishes were incubated at 25 °C, 37 °C and 45 °C in the dark for 7-14 days.

### Morphological characterization

Morphological characteristics of isolates were studied on PDA,OA, MEA, and SDA. After incubation, the diameters of the colonies on each agar media were measured. Colony color (obverse and reverse sides) and the degree of sporulation were also observed. Appropriate keys were used for the phenotypic identification of the isolated fungi [18] Mounted needle was used to pick ascomata, which were placed on slides using lactophenol cotton blue as the mounting medium and then examined under Olympus microscope (Olympus Optical Co., Ltd., Tokyo, Japan) connected with digital camera (KOPTIC Korea Optics, Seoul, Korea) `

### DNA extraction and phylogenetic identification

Genomic DNA was extracted from a fresh culture grown on MEA plates using the cetyltrimethyl ammonium bromide (CTAB) protocol of Möller et al. [41]. The ribosomal DNA (rDNA) internal transcribed spacer (ITS) gene and partial sequences of the actin(ACT1), β-tubulin(tub2), DNA-dependent RNA polymerase II largest subunit (RPB1) and second largest subunit (RPB2). and elongation factor 1α (TEF1) genes, as well as the 18S rDNA gene (small subunit [SSU]) and 28S rDNA gene (large subunit [LSU]), were amplified and sequenced. Primers used for amplification and sequencing are according to de Hoog et al. [37]. The PCR conditions were the same as those described by Wang et al. [18].

### Alignment and phylogenetic analyses

Consensus sequences from forward and reverse primers were generated using Seqman assembly programme from Lasergene software package (DNASTAR, Madison, WI, U.S.A.). These sequences were compared to the GenBank sequence database using nucleotide BLAST tool. Besides the sequences generated in this study, additional 25 sequences were retrieved from GenBank, along with the outgroup CBS24975 *Collariella gracills* (figure-5).

**Figure 5:**
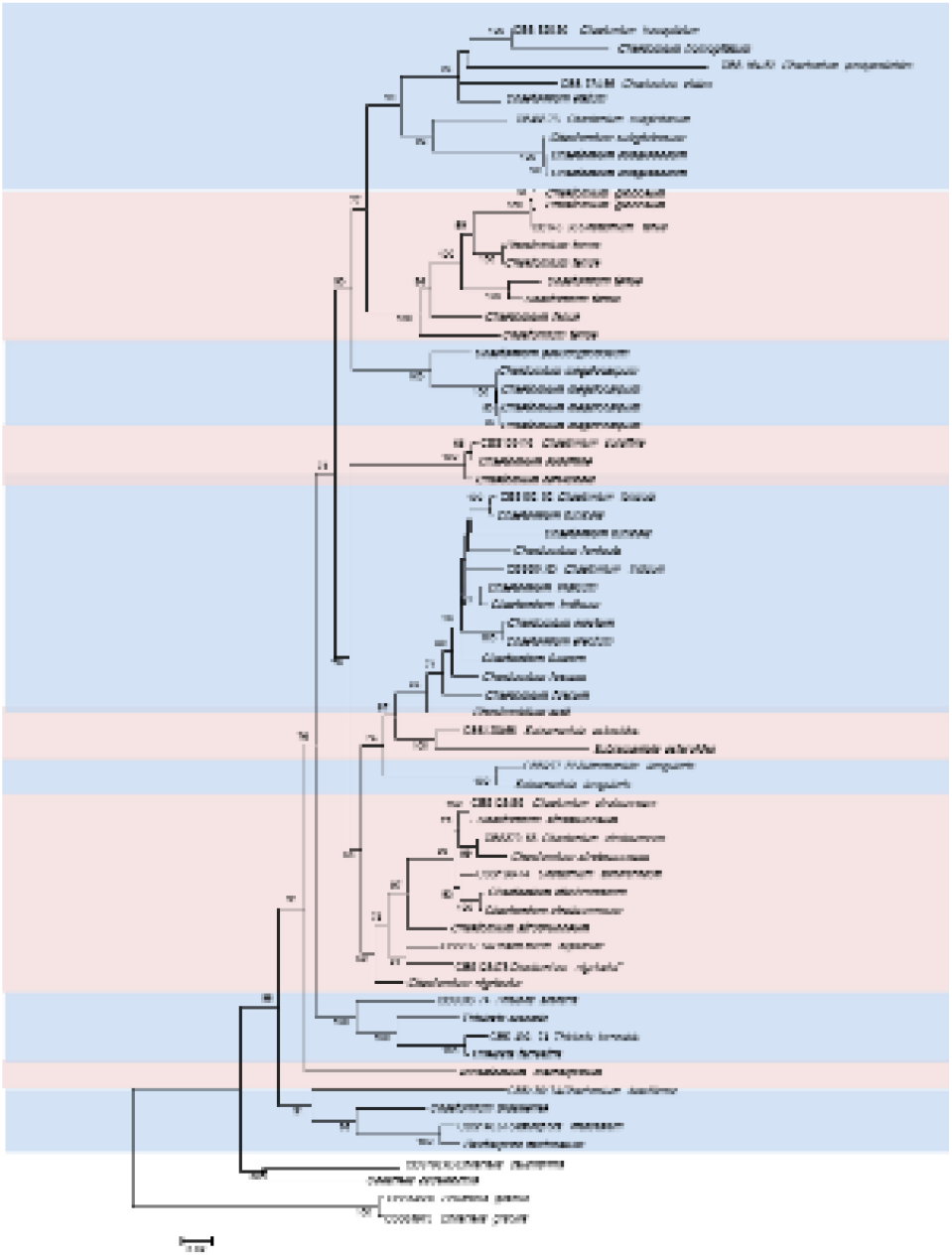
Phylogenetic tree of Chaetomiaceae family with closely related taxa from GenBank and their accession numbers generated from maximum likelihood (ML) analysis based on the combined dataset of ITS, partial LSU, tub2 and partial rpb2 sequences. with CBS24975 *Collariella gracills* as outgroup. Bootstrap values > 50% (1000 replicates).

The ITS region was used to initially screen the collection of fungi from the environments in order to select members of the Chaetomiaceae. The tub2 gene region was used to recognize the species diversity within the indoor Chaetomiaceae isolates. The phylogenetic placement of the environments isolates was determined using four loci (ITS, partial LSU, tub2 and partial rpb2) on the basis of the evaluation in a previous study [18], and representatives of related species and genera in the Chaetomiaceae were included as references from the gene bank in the final phylogenetic analyses.

Phylogenetic analyses were based on Bayesian inference (BI), Maximum Likelihood (ML) and Maximum Parsimony (MP) as described previously [18]. Gaps and missing data were deleted. For BI, the best evolutionary model for each locus was determined using MrModeltest v. 2.0 [42]. Confidence values were assessed from 1000 bootstrap replicates of the original data. Bootstrap values below 50% were removed from tree.

## RESULTS

### Geology of the study area

The characteristic of the soil from collection area was divided geographically into two categories. These are the sandy soils of the northern and west central areas in desert or semis desert ecozones, the clay soils of the central region in savanna ecozone (figure-1).

### Isolate

Chaetomiaceae were successfully isolated from different Sudanese environment. A total of 119 isolates (Table-1 and figure-5 in bold font) were identified as members of Chaetomiaceae after the ITS sequencing combined with an examination of the macro- and micromorphology. Out of 63 Chaetomium strains obtained from soil, animal dung and water samples, 25 were obtained from soil, 22 from animal dung and 16 from water.

**Table 1.**
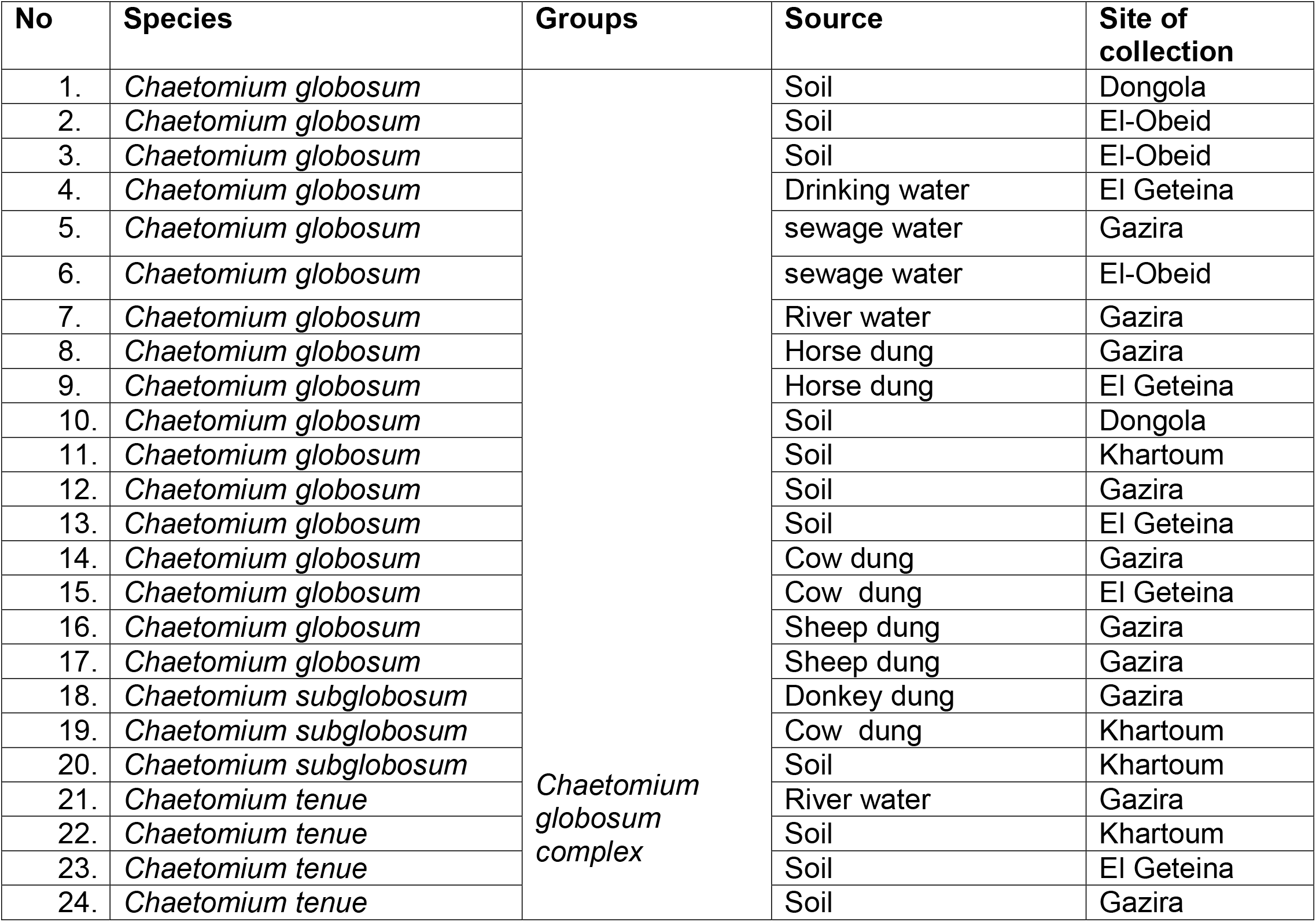
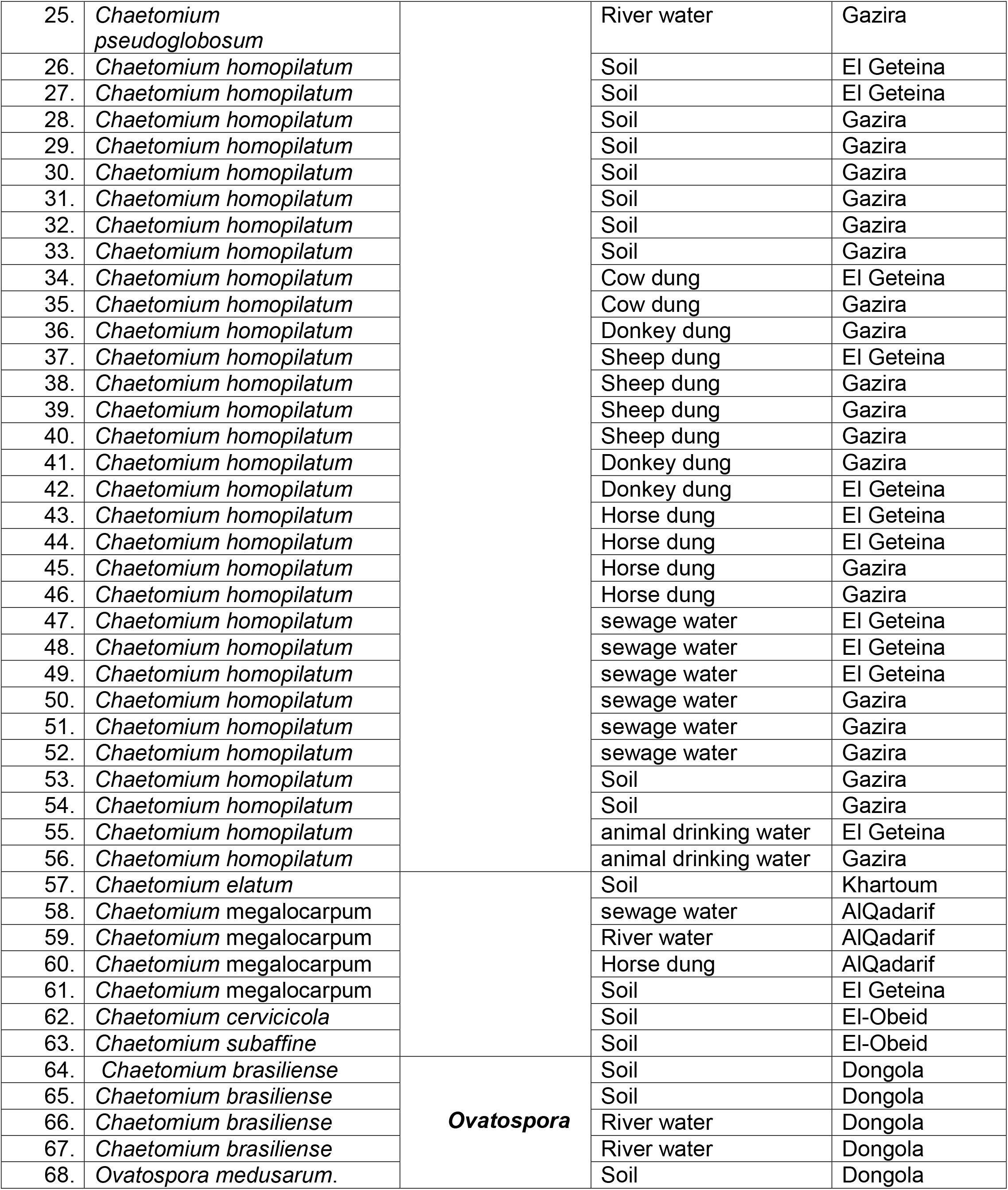
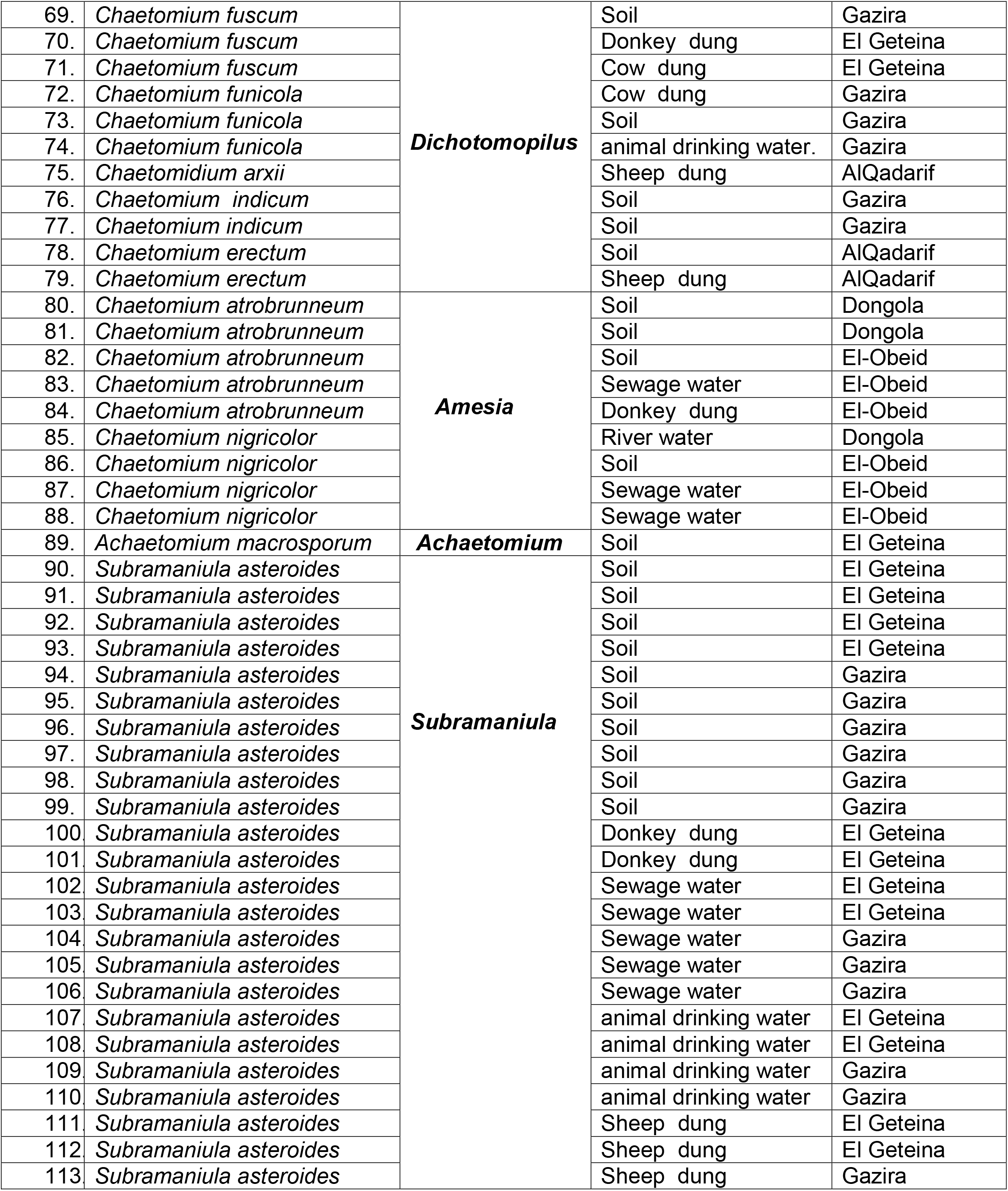
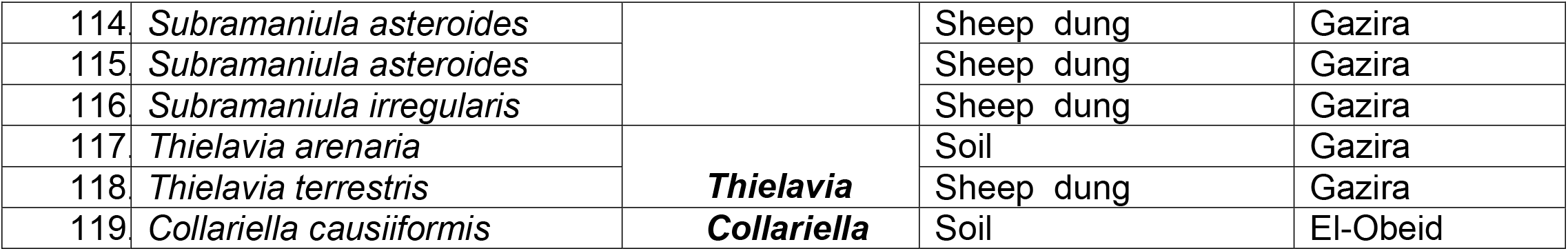
A list of all the strains isolated in this study

56 additional strains isolated from other genus within the Chaetomiaceae family, such as (*Amesia, Collariella, Ovatospora, Subramaniula* and *Thielavia*) were recorded for the first time in Sudan. Namely, *Amesia nigricolor, Collariella causiiformis, Ovatospora medusarum, Subramaniula asteroids, Subramaniula irregularis Thielavia arenaria* and *Thielavia terrestris*.

### Phylogeny

The phylogenetic relationships of Chaetomium species were studied using combined sequences of four-locus alignment (ITS, partial LSU, tub2 and partial rpb2) on the basis of a dataset consisting of 25 representative indoor isolates and representative isolates of related genera and species (figure-5)`. The concatenated alignment consisted of 3 570 characters (including alignment gaps).

The concatenated phylogenetic analyses revealed the *Chaetomium globosum* species complex and seven other monophyletic clades. In the backbone tree, most species can be clearly distinguished from each other, and the species clearly grouped in12 phylogenetic groups which showed a certain correspondence with their morphology profiles (figure-5). Our phylogenetic analyses also showed that the 119 strains isolated from Sudan in this study belonged to 22 species. In our collection, 52.9% of the indoor isolates with nine species cluster with members of the *Chaetomium globosum* species complex, included : *Chaetomium* globosum, *Chaetomium subglobosum, Chaetomium pseudoglobosum, Chaetomium elatum, Chaetomium tenue, Chaetomium homopilatum, Chaetomium subaffine, Chaetomium* megalocarpum, *Chaetomium cervicicola*. The other indoor species fall into seven known genera were supported: *Achaetomium* including the type species *Achaetomium. macrosporum*: Amesia with four indoor species *Amesia nigricolor* (=*Chaetomium nigricolor*) five species of *Amesia atrobrunneum, (=Chaetomium atrobrunneum)*, Collariella with one indoor species *Collariella causiiformis*, and Ovatospora with one indoor species *Ovatospora medusarum* and four species of *Ovatospora brasiliense* (=*Chaetomium brasiliense)*. Two isolates *Thielavia arenaria* and *Thielavia terrestris*. Three species of Dichotomopilus. funicola(=*Chaetomium funicola)* and *Dichotomopilus fuscum (=Chaetomium fuscum)*, Tow species of *Dichotomopilus indicum*,(=*Chaetomium indicum*) and *Dichotomopilus erectum* (=*Chaetomium erectum*), one species of *Dichotomopilus arxii* (=*Chaetomidium arxii)* Subramaniula is including two indoor species *Subramaniula asteroids, Subramaniula irregularis*.

According to this study, *Chaetomium homopilatum* is the most abundant Chaetomiaceae indoor species (31/119), followed by *Subramaniula asteroids* (27/119) and *Chaetomium globosum* (17/119).

To our knowledge all these species were recorded for the first time in Sudan and the morphological diversity of some indoor Chaetomiaceae are described below:

## Taxonomy

### Achaetomium macrosporum

*Achaetomium macrosporum* J.N. Rai, K. Wadhwani & J.P.Tewari, *Indian Phytopath*. 23 : 54 (1970).

*Achaetomiella macrospora* (Rai *et al*.) v. Arx, *Proc. Konink. Nederl. Akad. Wet. Amsterdam,ser*, C, 76: 292 (1973).

#### Microscopy

Hyphae 1-6 pm diameter, hyaline to mid brown, septate, frequently branched, sometimes nodulated, the older hyphae with a conspicuous brown gelatinous coating up to 2,um thick, sometimes producing reddish soluble exudates. Ascomata composed of dark brown textura intricata with irregular hyphae to 3um diameter, sometimes with an inner layer of dark brown textura epidermoidea with cells 3-8 um diameter. 140-290 × 110-210, um, ellipsoidal to pyriform, *Asci 55-80* x 12-19 um, clavate, fairly long-stalked, very thin-walled, without apical structures, evanescent, 8-spored. *Ascospores* arranged biseriately, 16.5-21.5 × 10-13.5 x *9-11* um, limoniform, flattened on two opposite lateral faces, sometimes curved along the longitudinal axis, the apices sometimes umbonate, dark chocolate brown, aseptate, rather thick walled, with a single apical germ pore 0.5-0.75um diameter.

#### Culture characteristic

colonies on MEA at 25° 70-75 mm diameter after 7 days Submerged mycelium hyaline, inconspicuous. Aerial mycelium very sparse, hyaline, lanose. Ascomata developing after about two weeks, covered in yellowish-green hypha-like hairs, scattered over the plate. On OA at *25°* colonies 75-80 mm diameter after 7 days Submerged mycelium obscured. Aerial mycelium copious, hyaline, floccose. Ascomata developing after about 2 weeks, scattered over the agar surface, greenish yellow, eventually turning dark brown. On PDA at 25° colonies 70-75 mm diam after 7 days. Submerged mycelium inconspicuous, hyaline. Aerial mycelium hyaline, sparse, lanose. Ascomata developing abundantly after about 2 weeks, greenish yellow to beige, scattered over the agar surface, eventually turning dark brown.

#### Material examined

This fungus was isolated from one soil samples taken from El - Geteina.

### Amesia

*Type species*: *Amesia atrobrunnea* (=*Cheatomium atrobrunneum*) (Ames) X. Wei Wang & Samson Stud. Mycol. 84: 158 (2016a).

#### Microscopy

Mycelium composed of hyaline to subhyaline, septate, smooth hyphae, 9.0−9.4 μm diameter. Superficial, ostiolate ovate, or subglobose ascomata with dimensions of 75-165 μm × 70-140 μm, black in color. The wall of the ascomata was black to dark brown in color. The hairs were flexuous, smooth, and septate and were 2.3-3.5 μm in diameter near the base. Had eight ascospores and clavate, evanescent, 9-22 μm-long asci. The ascospores were fusiform or elongate and turned dark brown when they matured7.5-10× 4-5.5μm.

#### Culture characteristics

Colonies on OA with an entire edge, about 41–47 mm diameter in 7 days at 25 °C, fuscous black to black, with fawn to greyish sepia exudates diffusing into the medium, reverse greyish sepia to black. Colonies on PDA with an entire edge, about 39–45 mm diameter in 7 days at 25 °C, translucent, vinaceous buff and floccose due to ascomata mixed with aerial hyphae, without coloured exudates; reverse uncoloured. Colonies on MEA with an entire edge, about 36–42 mm diameter in 7 days at 25 °C, with greyish white and floccose mycelium, looser and white to pale olivaceous buff texture in the central part due to ascomata mixed with aerial hyphae, reverse ochraceous.

#### Material examined

This fungus was isolated from three soil samples, one sewage water sample and one donkey dung sample taken from El-Obeid and Dongola state

### Amesia nigricolor

(=*Cheatomium nigricolor*) (Ames) X. Wei Wang & Samson, comb..Basionym: Chaetomium nigricolor Ames, Mycologia 42: 654.1950..

#### Microscopy

Hyphae hyaline, septate, smooth hyphae, 9.5−9.9 μm diameter Ascomata superficial, ostiolate, olivaceous grey in color. Ascomatal hairs, subglobose to ovate, 140–300 μm high, 100–255 μm diam. Ascomatal wall brown. Terminal hairs undulate to loosely coiled with erect or flexuous lower part, conspicuously rough (granulate), greyish sepia to brown, septate, 3–4.5 μm diameter in the undulate or coiled upper portion. Lateral hairs flexuous, un-dulate or apically circinate. Asci fasciculate, clavate to fusiform, spore-bearing part 13.5– 21 × 7.5–10.5 μm, stalks 6–11.5 μm long, with 8 irregularly-arranged ascospores, evanescent. Asco-spores olivaceous brown when mature, ovate, (5.5–) 6–7(–7.5) × 4–5(–5.5) μm, with an apical germ pore at the attenuated end.

#### Culture characteristics

Colonies on OA with entire edge, about 42–48 mm diameter in 7days at 25 °C, with sparse white aerial hyphae, with ochraceous to fulvous exudates diffusing into the medium, reverse pale luteous to amber. Colonies on PDA with entire edge, about 35–41 mm diameter in 7 days at 25 °C, with sparse white to pale buff aerial hyphae; reverse uncoloured. Colonies on MEA with entire edge, about 47–53 mm diameter in 7 days at 25 °C, with white floccose aerial mycelium, reverse sienna with pale edge.

#### Material examined

This fungus was isolated from one soil samples and one river water sample and two sewage water samples taken from El-Obeid and Dongola state

### Chaetomium

*Type species*: ***Chaetomium globosum*** *Chaetomium globosum* Kunze, Mykol. Hefte 1: 16. 1817

#### Microscopy

Hyphae hyaline, partially converting to dark brown or black, Ascomata superficial, ostiolate, greenish olivaceous, or slightly dark olivaceous buff to grey, or dull green in color. Ascomatal hairs sulphur yellow lower parts or appear as sulphur yellow to yellow, sub-globose, ovate or obovate, 140–270 μm high, 100– 240 μm diameter. Ascomatal wall brown.. Terminal hairs finely warty, brown, undulate to loosely coiled with erect or flexuous lower part, tapering and fading towards the tips, 2–5 μm diameter near the base. Lateral hairs brown, flexuous, fading and tapering towards the tips. Asci fasciculate, fusiform or clavate, spore-bearing part 19–38 × 12–17 μm, stalks 22–48 μm long with 8 irregularly-arranged ascospores, evanescent. Ascospores brown when mature, limoniform, usually biapiculate, bilaterally flattened, 8.5–11(–12) × 7–8.5(–9.5) × 5.5–7 μm.

#### Culture characteristics

Colonies on OA with an entire edge, spreading rapidly, over 70 mm diameter in 7days at 25 °C, with sparse white to olivaceous buff aerial hyphae when young, then without aerial hyphae and becoming citrine green, yellow, greenish olivaceous to grey olivaceous owing to the aggregation of ascomata, with olivaceous to olivaceous grey exudates diffusing into the medium; reverse grey olivaceous to olivaceous. Colonies on PDA translucent, usually with a more or less lobate edge, about 39–48 mm diameter in 7 days at 25 °C, without or with very sparse and floccose, pale yellow aerial hyphae, without coloured exudates; reverse uncoloured. Colonies on MEA with an entire edge, spreading rapidly, over 70 mm diameter in 7 days at 25 °C, honey to olivaceous buff in color, reverse fulvous, orange to sienna with exudates diffusing into the medium (figure-6)..

**Figure 6:**
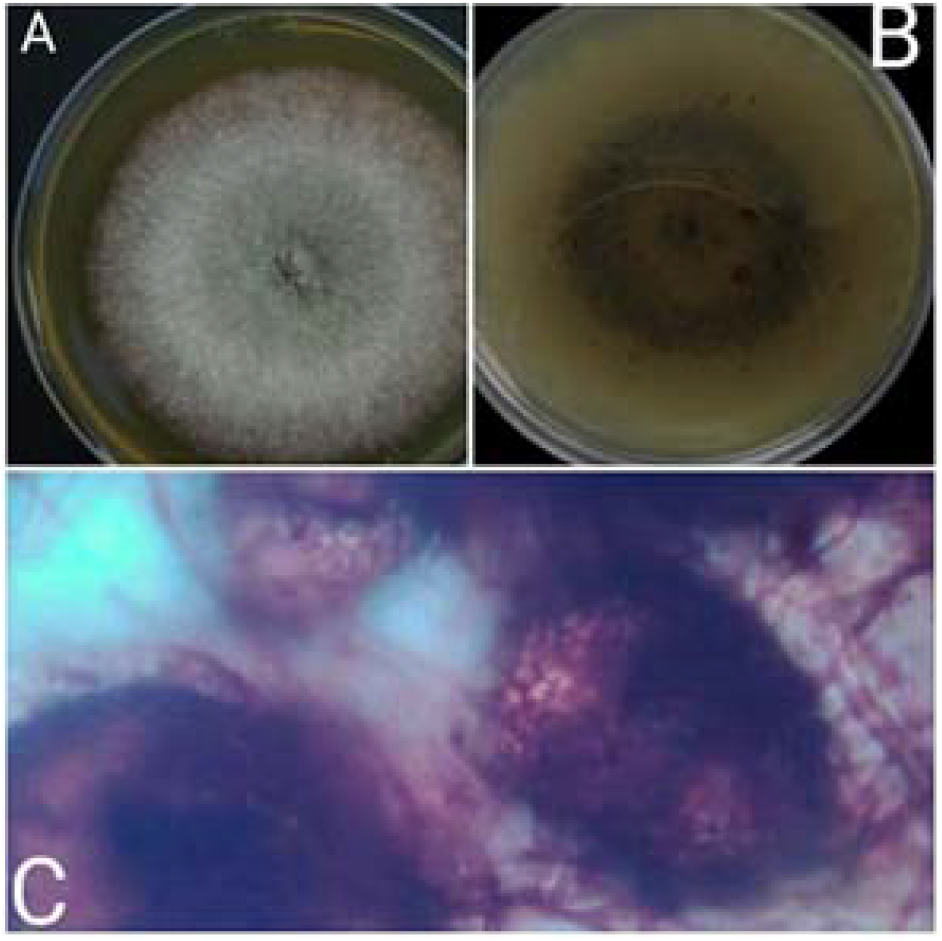
*Chaetomium globosum* : A., Colonies on PDA after 7 days incubation from Surface side and B from reverse side C. Mature ascomata

#### Material examined

This fungus was isolated from seven soil samples, two sewage water samples, one drinking water sample, one river water sample, two cow dung samples, two Sheep dung samples, and two horse dung samples taken from El-Obeid, Dongola, El Geteina, Gazira and Khartoum state.

### Chaetomium elatum

*Chaetomium elatum* Kunze, Deutsche Schw€amme 8: 3, No. 184. 1818.

#### Microscopy

Hyphae hyaline, partially converting to dark brown or black. Ascomata superficial, ostiolate often covered by sparse white or buff aerial hyphae, grey olivaceous to isabelline, globose or obovate, 280–440 μm high, 255–380 μm diameter. Ascomatal wall brown. Terminal hairs verrucose or warty, dark brown, tapering and fading towards the tips, erect or flexuous in the lower part, 3–5 μm diameter near the base, erect to flexuous terminal branches taping and fading towards the tips. Lateral hairs brown, erect or flexuous, occasionally undulate, tapering towards the tips. Asci fasciculate, clavate or fusiform, spore-bearing part 36–49 × 12.5–16.5 μm, stalks 24–55 μm long, with 8 biseriate ascospores, evanescent. Asco-spores brown when mature, limoniform, biapiculate or umbonate, bilaterally flattened, (10–)11.5–13.5(–16) × (8–) 8.5–10(–11) × (6.5–)7–8 μm..

#### Culture characteristics

Colonies on OA with entire edge and spreading rapidly, over 70 mm diameter in 7 days at 25 °C, with sparse, honey to greenish olivaceous aerial hyphae, without coloured exudates when young, or with greyish yellow-green, olivaceous buff to honey exudates diffusing into the medium, reverse honey to greenish olivaceous. Colonies on PDA pale luteous to luteous, with irregularly deep lobate edge, about 52–61 mm diameter in 7 days at 25 °C, aerial hyphae sparse and honey, with luteous exudates diffusing into the medium; reverse pale luteous to amber. Colonies on MEA forming pale honey, citrine green to citrine and floccose mycelium with entire edge, about 58–66 mm diameter in 7 days at 25 °C, reverse citrine green to citrine.

#### Material examined

This fungus was isolated from one soil sample taken from Khartoum state

### Chaetomium homopilatum

Omvik, Mycologia 47(5): 749.1955.

Ascomata 150–206 × 84–120 μm, superficial, membranous, solitary, hairy, ovoid, brown; terminal hairs forming a dense tuft around the ostiole, up to 4 μm wide at base, septate, verrucose, tapering at apex, light brown; lateral hairs shorter, up to 4 μm wide at base and randomly distributed over the perithecium, light brown. Peridium composed of angular cells. Paraphyses not observed. Asci not observed. Ascospores 6–7 × 4.5–6 μm, discharged as a cirrus, smooth, aseptate, rather thin-walled, broadly oval, apiculate at both ends with a brown germ pore (figure-7).

**Figure 7:**
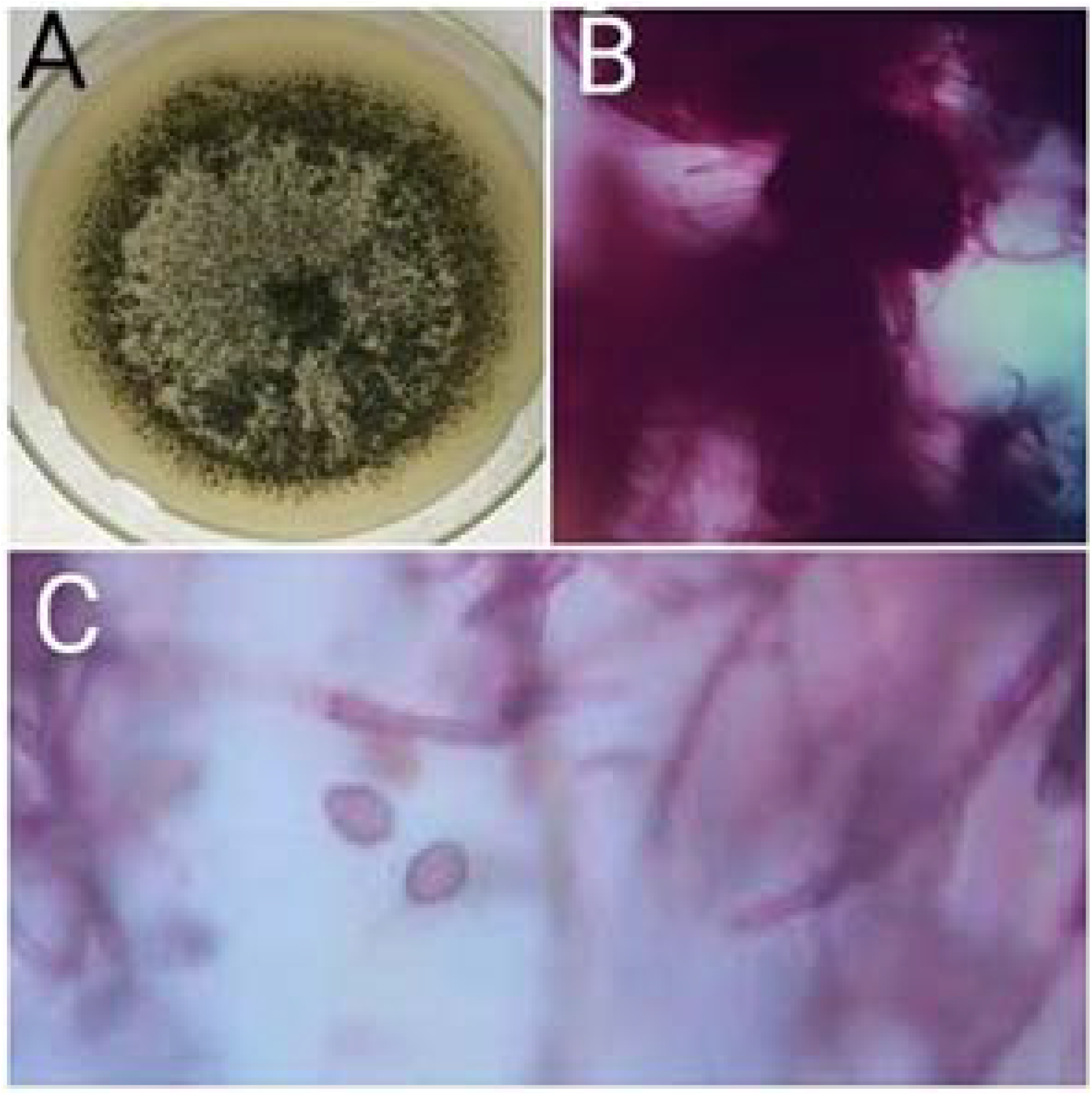
*Chaetomium homopilatum*: A., Colonies on PDA after 7 days incubation from Surface side; B Mature ascomata, showing terminal coiled terminal hairs; C. Asci

#### Material examined

This fungus was isolated from ten soil samples, six sewage water samples, two animal drinking water, three donkey samples, two cow dung samples, four Sheep dung samples, and four horse dung samples taken from El Geteina and Gazira state

### Chaetomium subaffine

*Chaetomium subaffine* Sergeeva, Not. Syst. sect. Crypt. Inst. Bot. Acad. Sci. 14: 148. 1961.

#### Microscopy

Hyphae hyaline, partially converting to dark brown or black. Superficial, dark, obovate, or ovate ascomata with dimensions of 220-410 μm × 180-340 μm, olivaceous or dark. The ascomatal wall was brown. The hairs were erect to flexuous, septate, verrucose, generally unbranched and tapering towards the tips, and 2.6-4.2 μm in diameter near the base. Had eight ascospores within clavate and sometimes slightly fusiform evanescent, 17-36 μm-long asci. The ascospores were bilaterally flattened limoniform, and usually biapiculate, when mature become brown, 11.5-13.5 (−14) μm × 8-10 μm × 6-8.2 μm in size.

#### Colony morphology

Colonies exhibited good growth and matured within 7-10 days on OA medium at 25 °C. They were nearly 45-50 mm in diameter, black, and had abundant white aerial hyphae. They were uncolored in reverse.

#### Material examined

This fungus was isolated from one soil sample taken from El-Obeid.

### Collariella

#### Collariella causiiformis

Collariella causiiformis (Ames) X. Wei Wang & Samson, comb. Basionym: Chaetomium causiiforme Mycologia 41: 644. 1949.

##### Microscopy

Hyphae hyaline, partially converting to dark brown or black. Ascomata superficial, usually forming a layer on the surface of OA medium. Ascomatal hairs hazel to olivaceous in reflected light due to ascomatal hairs, globose or subglobose, ostiolate, 70–140 μm high (including the darkened collar), 60–120 μm diameter, apically black due to a darkened collar in 10–24 μm high and 32–65 μm wide around the ostiolar pore. Ascomatal wall translucent, ochraceous, textura angularis in surface view. Asci fasciculate, fusiform, sometimes clavate, spore-bearing portion 15–22 × 8–13 μm, stalks 6–13 μm long, with 8 irregularly-arranged ascospores, evanescent. Asco-spores olivaceous when mature, broad limoniform or ovate, bilaterally flattened, (5–)5.5–6.5 × (4.5–)5–5.5(–6) × (3.5–) 4–5 μm.

##### Culture characteristics

Colonies on OA translucent at the beginning, with entire edge, 27–33 mm diameter in 7 days at 25 °C, without aerial hyphae, without coloured exudates, reverse uncoloured. Colonies on PDA translucent, with entire edge and lobate and relatively thicker texture in the centre, about 24–30 mm diameter in 7 days at 25 °C, without aerial hyphae, without coloured exudates; reverse uncoloured. Colonies on MEA translucent and membranous, with entire edge, about 20–26 mm diameter in 7 days at 25 °C, forming radiating and luteous or fulvous furrows, non-sporulating; reverse uncoloured.

##### Material examined

This fungus was isolated from one soil sample taken from El-Obeid.

### Dichotomopilus

#### Dichotomopilus funicola

*Dichotomopilus funicola (Chaetomium funicola)* (Cooke)

X. Wei Wang & Samson & Crous.

*Basionym*: *Chaetomium funicola* Cooke, Grevillea 1: 176. 1873.

*Synonym*: *Chaetomium cancroideum* Tschudy, Am. J. Bot. 24: 478. 1937.

##### Microscopy

Hyphae hyaline, partially converting to dark brown or black. *Ascomata* superficial, ostiolate, greenish olivaceous to grey olivaceous, subglobose to ellipsoid or ovate, 195–255 μm high, 180–240 μm diameter. *Ascomatal wall* composed of brown, irregular or elongate cells. *Terminal hairs* either to be wider, dark brown and erect, 4–5.5 μm diameter near the base, tapering and fading towards tips, dichotomously branched profusely at acute angles starting from the upper half part, punctulate; or thinner, luteous to brown, 2–3.5 μm diameter near the base, dichotomously branched at wide to acute angles starting near the base; the terminal branches relatively short, erect, incurved or reflexed, often in dense clusters. *Lateral hairs* simply branched or seta-like, tapering and fading towards tips. *Asci* fasciculate, with 8 irregularly-arranged ascospores, clavate, pyriform to ovate, spore-bearing portion 11–18 × 7–11.5 μm, stalks 6–10 μm long, evanescent quickly. *Ascospores* olivaceous when mature, ovate to slightly elongate ovate, bilaterally flattened, 5–6 × 3.5–4.8 × 3–3.8 μm.

##### Culture characteristics

Colonies on OA with an entire edge, spreading rapidly, over 70 mm diameter in 7 days at 25 °C, with pale yellow, sparse and floccose aerial hyphae, producing greenish olivaceous to citrine exudates diffusing into the medium; reverse ochraceous to cinnamon. Colonies on PDA olivaceous buff to greenish olivaceous with irregularly crenated edge, spreading rapidly, over 70 mm diameter in 7 days at 25 °C, without coloured exudates; reverse olivaceous buff to greenish olivaceous. Colonies on MEA with entire edge, about 57–63 mm diameter in 7 days at 25 °C, forming profuse and pale yellow mycelium and then greenish olivaceous without coloured exudates, reverse ochreceous.

##### Material examined

This fungus was isolated from one soil sample, one animal drinking water sample and one cow dung sample taken from Gazira state

### Dichotomopilus indicus

#### Dichotomopilus indicus

(*Chaetomium indicum*) (Corda) X. Wei Wang & Samson.

*Basionym*: *Chaetomium indicum* Corda, Icon. Fung. 4: 38. 1840.

##### Microscopy

Hyphae hyaline, partially converting to dark brown or black. *Ascomata* superficial, ostiolate, greenish olivaceous to grey olivaceous in reflected light, subglobose or ovate, 160–310 μm high, 140– 275 μm diameter. *Ascomatal wall* composed of brown, irregular or elongate cells. *Terminal hairs* composed of one type: dark brown and erect, 3–5.5 μm diameter near the base, dichotomously branched at acute to wide angles starting from the upper half part, verrucose, tapering and fading towards tips. *Lateral hairs* unbranched, seta-like, tapering and fading towards tips. *Asci* fasciculate, with 8 irregularly-arranged ascospores visible, clavate, broadly clavate, fusiform to pyriform, spore-bearing portion 11–25.5×6.5– 13.5 μm, stalks 6–15 μm long, evanescent quickly. *Ascospores* olivaceous when mature, ovate to slightly elongate ovate, bilaterally flattened, 5.5–6.5 × 3.5–4.5 × 2.5–3 μm.

##### Culture characteristics

Colonies on OA with entire edge, about 20–48 mm diameter in 7 days at 25 °C, with sparse buff, pale yellow to pale primrose aerial hyphae, without coloured exudates at the beginning, and then producing pale luteous, amber, ochraceous to cinnamon exudates diffusing into the medium, reverse buff, pale luteous, luteous, ochraceous to cinnamon. Colonies on PDA usually with entire, slightly undulate, or irregularly crenated edge, about 25–54 mm diameter in 7 days at 25 °C, olivaceous buff, pale yellow to greenish olivaceous without coloured exudates; reverse olivaceous buff, pale yellow to greenish olivaceous. Colonies on MEA with entire or slightly lobate edge, about 36–53 mm diameter in 7 days at 25 °C, forming thick yellowish white to pale yellow mycelium, sometimes with radiating furrows, reverse pale luteous to ochraceous.

##### Material examined

This fungus was isolated from two soil samples taken from Gazira state

***Ovatospora*** X. Wei Wang, Samson & Crous.

#### Ovatospora brasiliensis

*Ovatospora brasiliensis* (Batista & Pontual) Wang & Samson, comb. nov., Stud. Mycol. 84: 207 (2016a).

Basionym: *Chaetomium brasiliense* Batista & Pontual, Bol. Agr. Com. Pernambuco 15: 70. 1948.

#### Microscopy

Hyphae hyaline, partially converting to dark brown or black. Superficial, pale gray, subglobose, or globose ascomata 85-135 μm × 72-115 μm in size, pale olivaceous gray in reverse light. The ascomatal wall was brown. The hairs were loosely coiled or undulate in the upper part and erect in the lower part and obviously rough, septate, grayish to brown in color, and 2-3.7 μm in diameter in the upper part. The asci cylindrical, evanescent, 34-42 μm-long with eight ascospores. The ascospores were ovate, bilaterally flattened, turned brown when mature, 6.5-7.7 μm × 5.3-6.2 μm × 5-6.7 μm in size.

#### Culture characteristics

Colonies were matured within 7 days on OA medium at 25 °C. They were nearly 40-50 mm in diameter, pale gray to pale olivaceous gray, and black in reverse. Colonies on PDA translucent, with entire edge, about 36–42 mm diameter in 7 days at 25 °C, with olivaceous buff and floccose aerial hyphae mixed with ascomata, without coloured exudates; reverse olivaceous or olivaceous grey. Colonies on MEA with slightly undulate edge, about 32–38 mm diameter in 7 days at 25 °C, with floccose, concentric and buff to white aerial mycelium, with pigmented hyphae immersed in medium, without coloured exudates, reverse black with pale edge.

#### Material examined

The examined materials were isolated from two soil samples and two river water samples collected from Dongola state.

#### Subramaniula

Arx, Proc. Indian Acad. Sci., Plant Sci. 94: 344. 1985. ***Subramaniula asteroides*** :*Subramaniula asteroides* S.A. Ahmed, Z. U. Khan, X. Wang & de Hoog, sp. nov. Fungal Diversity (2016) 76:11–26.

#### Microscopy

Hyphae broad, septate, hyaline, turning dark brown with age, verruculose; part of the hyphae convert to dark brown or black, thick walled chlamydospore-like structures. Cellular clumps irregular, black 58–100×44–71 μm, consisting of aggregates of dark brown, thick-walled cells 7–12 × 7–9 μm. Conidia hyaline 2–4×1.5– 2.0 μm, unicellular, obovoidal or ellipsoidal

#### Culture characteristics

Colonies on OA medium at 25 °C, felty, yellowish green or fuscous at the centre becoming faint toward the margin; some isolates showing dark, waxy colonies covered with black bulbils with age. Colonies on MEA radially folded, yellow green, turning dark greyish green with age; reverse dark grey.

#### Material examined

This fungus was isolated from ten soil samples, five sewage water samples, four animal drinking water samples, six sheep dung samples and two donkey dung samples taken from El Geteina and Gazira state.

### Thielavia

#### Thielavia terrestris

Thermothielavioides terrestris (Apinis) X. Wei Wang & Hou-braken, comb. nov. Basionym: Allescheria terrestris Apinis, Nova Hedwigia 5: 68. 1963. Synonym: Thielavia terrestris (Apinis) Malloch & Cain, Canad. J. Bot. 50: 66. 1972. **Microscopy:** Ascomata superficial or covered by aerial mycelium, solitary to aggregated, non-ostiolate, lead black in reflected light, globose or subglobose, 80– 270 μm diameter. Ascomatal wall brown, non-translucent or semi-translucent, composed of irregular, angular or elongated cells. Ascomatal hairs brown, septate, flexuous, verrucose, tapering and fading to hyaline towards tips, 2–3.5 μm diameter near base. Asci ellipsoidal to ovoid, spore-bearing part 13.5–20.5 × 6–8 μm, with stalks 4.5–11.5 μm long, containing eight biseriate or irregularly arranged ascospores, evanescent. Ascospores 1-celled, olivaceous brown when mature, smooth, ellipsoidal or ovoid, 4–6.5(–7) × (3–)3.5–4.5(–5) μm. Conidiogenous cells arising laterally from aerial hyphae, hyaline, phialidic, occasionally branched, (6.5–)9–24(–33) × (1–)1.5–3.5 μm. Conidia in basipetal chains, hyaline, aseptate, smooth, obovoid to clavate, usually with a truncated base and a rounded apex, (3–)3.5–5(–5.5) × 1.5–2.5(–3) μm.

#### Culture characteristics

On OA with an entire edge, 53–59 mm diameter in 7 days at 25 °C, texture thick floccose, obverse white to pale smoke grey, or rosy buff to hazel, reverse cinnamon to hazel. On PDA similar to those on OA. On MEA with an entire edge, 39–45 mm diameter in 7 days at 25 °C, texture thick floccose, obverse white, reverse fawn. On PDA with an entire edge, 41–47 mm diameter in 7 days at 25 °C, with sparse aerial mycelium, producing ascomata when growing on cellophane membrane covering the surface of the medium, obverse white to smoke grey, reverse smoke grey.

#### Material examined

This fungus was isolated from isolated from one sheep dung sample collected from Gazira state.

## DISCUSSION

Members of the family *Chaetomiaceae* have been globally reported as being are capable of colonizing various substrates and are well-known for their ability to degrade cellulose and to produce a variety of bioactive metabolites [12–15]. They distribute widely in the diverse climates from the tropics to temperate regions. In spite of huge differences among geographical and ecological factors in the distribution area, the taxonomic position of *Chaetomiaceae* is uncertain due to limited number of morphological and developmental characters.

This family is characterized by the formation of ascospores in asci inside perithecia. Phenotypic identification of species in *Chaetomiaceae* mostly depends on the shape of ascomatal hairs, ascoma pigmentation and ascospore shape [4, 9].

In this study, during a survey of Chaetomium and Chaetomium like fungi in soils, dung and water from various places in Sudan, several isolates were collected and identified based on morphological and molecular analysis.

Like many sub-Saharan African countries, including Sudan, have considerably diverse soils, such as clay in the east-central area and desert in the west and north, all with variable climatic conditions [43,44] A range of isolates from these soil types with different state were noticed.

Our result showed that the dominant species in this study was *Chaetomium homopilatum* and *Chaetomium globosum* in both state, Gazira and El Geteina. While the genus Subramaniula with *Subramaniula asteroids* species was found to be the second most common isolate in the indoor environments. Similar to our result; in previous study *Chaetomium homopilatum* was found on different substrates, such as dung, leaves, wood, and soil in terrestrial habitats [10]. In Brazil, this species was found on *Saccharum officinarum* [45] and from freshwater habitats [46].

The presence of the of these fungi in the dung, soil and all water types is due to the fact that The Gezira and El Geteina state local villages are characterized by an abundance of cattle, goats, sheep, horse and donkeys [47] Most inhabitants live on cattle and sheep husbandry and agriculture [48] (figure-1 D and J).they are built their house from muddy bricks and cement material made of soil, animals dungs and parts of the bushed in the villages that are rich of organisms [47].

The role of mammal dung and dung-enriched soil is one of the prime ecological niches for the species in *Chaetomiaceae* [35]. Multiple Chaetomium and Thielavia species have been isolated in East Africa from different kinds of dung, ranging from cow and horse to more exotic types of dung such as of elephant and wildebeest [49]. Thielavia was known as the second largest genus in the family [18]. The genus Thielavia is morphologically defined by having non-ostiolate ascomata with a thin peridium composed of textura epidermoidea, and smooth, single-celled, pigmented ascospores with one germ pore [18].

Contamination soil and dung with *Chaetomium* is predisposing factor for opportunistic infection. It is recommended to use building material not containing animals dungs and soil, and if that proved to be mandatory for socioeconomic factors, then they should be burn properly to destroy these causative organisms.

It has been demonstrated that the presence of *Chaetomium. globosum* in the indoor environment is one of the important contributors to the development of symptoms of rhinitis, asthma and other health problems [18] This species is also the most common human pathogen mainly associated with onychomycosis [19–26] In addition, some other indoor chaetomium-like species were also reported in clinical cases, such as *Dichotomopilus. funicola* (=*Chaetomium funicola*)[50], *Ovatospora brasiliensis* (=*Chaetomium brasiliense*) [24], and *Amesia* atrobrunnea (=*Chaetomium atrobrunneum*) [30,37,51]. These species, which grow in our living and working environment, deserve more attention in future.

In our result *Chaetomium funicola* was isolated from cow dung, soil and animal drinking water. in literature *Chaetomium funicola* is a widespread species and has been found as an endophyte of grass [52], an opportunistic fungus causing cutaneous lesions [53], a plant pathogen [54], as a decomposer on plant debris in soil [55], and in dung of various animals [4]. Although *Chaetomium funicola* is common in terrestrial habitats, it has been previously reported from wood in fresh water from North America [56] and in a mangrove swamp from India [57].

Most members of the Chaetomiaceae lack anamorph sporulation, or some scattered, undiagnostic phialides are present at most [9]. Thus, if strains lose the ability to produce their elaborate ascomata, they cannot be recognized as a Chaetomium species by morphological means. Most of the environmental Chaetomium strains analyzed in this study produced ascomata in culture, except *Chaetomium cervicicola* remained sterile

The genus *Subramaniula* was described by von Arx (1985) [7] to mark the 60^th^ birthday of Professor C. V. Subramanian. Subramaniula was distinguished from Chaetomium by its urniform and nearly glabrous pale ascomata with wide apical openings surrounded by a hyaline collar [7,10]. Our result showed that *Subramaniula asteroids* is second common enviromental fungi isolated from soil, animal dung and water while only one species *Subramaniula irregularis was* isolated from sheep dung. Subramaniula thielavioides has been isolated from a dung sample from India, whereas. Subramaniula irregularis is only known from soil in South Africa [8,58].

*Achaetomium*, a genus in the Chaetomiaceae was described by Rai *et al*. (1964) for fungi which were similar to *Chaetomium* Kunze but which were ‘ devoid of hairy ornamentation. The separation of *Achaetomium* from *Chaetomium* was reported by Rai *et al*. (1970) [59] who described a six species of the genus *Achaetomium* from India [60–65] A further species were described from mangrove swamps in eastern India [57]. A further species were described from arid soil in Egypt[66] Japan [67,68] and Nepal [69] Then in our study from El Geteina soil A single species of *Achaetomium macrosporum* was isolated. The species placed in this genus are now known from a wide range of countries in Africa, Asia and Australasia. Now a day It comprises 10 species that produce similar cleistothecial ascomata with a membranous, mostly pilose, peridium. included, *Achaetomium globosum, Achaetomium thermophilum, Achaetomium cristalliferum, Achaetomium luteum Achaetomium umbonatum, Achaetomium lippiae, Achaetomium aegilopis, Achaetomium sphaerocarpum, Achaetomium thielavioides, Achaetomium macrosporum*. Although none of the species described by these authors have well-developed hairs typical of the genus *Chaetomium*, they all in fact have strongly tomentose ascomata. The genus can most easily be distinguished from *Chaetomium* by the colour of the ascospores, which are dark chocolate brown in *Achaetomium* and pale to mid brown or olivaceous in *Chaetomium*. In many species the colour does not develop fully until the ascospores are released from the fruit body, so care must be taken not to base observation of this character on immature specimens. The species of *Achaetomium* also appear to lack periphyses, whereas at least most species of *Chaetomium* possess them, and *Achaetomium* ascomata are thick-walled, and composed at least partially of loosely knit textura intricata, while in *Chaetomium* they are relatively thin-walled, and composed of well-ordered textura intricata or angularis.

The name of this genus *Ovatospora* came from the ovate to broadly ovate ascospores of all of its species [18]. This species was previously isolated from moist jute cloth and from dried freshwater fish, prawns, and shrimps [18,70]. To our knowledge, this is the first record of *Ovatospora brasiliensis* from desert soil From Dongla in this study. *Ovatospora. brasiliensis* is of medical interest considering that it was isolated from a patient with spinocellular carcinoma, which is a case of otitis externa [24]

These results implied an active differentiation of morphology within these species lineages. This makes it very complicated to identify these species only based on morphology. A molecular based identification will help to solve this problem. Several Chaeotmiaceae species share ITS sequences, and for routine identification, β-tubulin (tub2) is recommended. On the other hand, it needs to be noted that not all species are represented with tub2 sequences in the public databases and future studies should aim to complete this omission.

In conclusion, this study was the first study for the family Chaetomiaceae in Sudan that relied on molecular analysis. The species within this family having diverse morphological characteristics, so the identification of these fungi depending on morphological characters is not enough and must be supported by modern phylogenetic techniques. Sequence-based identification of fungal isolates is often considered to be the most reliable and accurate identification method.

## Data Availability

All data produced in the present study are available upon reasonable request to the authors

## Abbreviations used in this paper

(rDNA): The ribosomal DNA
ITS: internal transcribed spacer gene
BT2: the first β-tubulin locus
TUB: the second β-tubulin locus
ACT1: partial sequences of the actin
RPB1: DNA-dependent RNA polymerase II largest subunit
RPB2: second largest subunit
TEF1.: elongation factor 1α genes

## Declarations

## Acknowledgements

We would like to thank all health centers’ staff for their kind collaboration and assistance. And also great thanks to all participants contributed to this work.

## Authors’ contributions

NAM was provided conceptual framework for the project, participated in data collection and analysis, participated in the molecular performance and writing the manuscript

## Funding

Not applicable.

## Availability of data and materials

The datasets used and/or analyzed during the current study are available from the corresponding author on reasonable request

## Ethics approval and consent to participate

Not applicable.

## Consent to publish

Not applicable.

## Competing interests

The authors declare that they have no competing interests

## Notes

### Competing Interest Statement

The authors have declared no competing interest.

### Funding Statement

This study did not receive any funding

## References

1. Kunze G, Schmidt JK (1817). Mykologische Hefte, : 1. Germany, Leipzig. Lagace J, Cellier E. A case report of a mixed Chaetomium globosum/ Trichophyton mentagrophytes onychomycosis. Medical Mycology Case Reports 2012 ; 1: 76–78.

2. Corda ACJ (1840). Icones fungorum hucusque cognitorum. Vol. 4. Praha. Cunningham CW. Can three incongruency tests predict when data should be combined? Molecular Biology and Evolution 1997; 14: 733–740.

3. Chivers AH. A monograph of the genera Chaetomium and Ascotricha. Memoirs of the Torrey Botanical Club 1915;14: 155–240.

4. Ames LM. A monograph of the Chaetomiaceae. U.S. Army Research and Development 1963; Series 2. 125pp.

5. Carter A. A taxonomyic study of the ascomycete genus Chaetomium Kunze. Ph.D. dissertation. University of Toronto, Canada. 1982

6. Carter A. Three new species in the genus Chaetomium. Canadian Journal of Botany 1983;61: 2603–2607.

7. Von Arx JA. On Achaetomium and a new genus Subramaniula (Ascomycota). Proc Indiana Acad Sci 1985; 94:341–345.

8. Von Arx JA, Mukerji KG, Singh N. A new coprophilous ascomycete from India. Persoonia 1978; 10:144–146.

9. Von Arx JA, Dreyfuss M,Müller E. A revaluation of Chaetomium and Chaetomiaceae. Persoonia 1984; 12:169–179

10. Von Arx JA, Guarro J, Figueras MJ. The ascomycete genus Chaetomium. Beih Nova Hedwig 1986; 84:1–162.

11. Wang XW, Lombard L, Groenewald JZ, Li J, Videira SI, Samson RA, et al. Phylogenetic reassessment of the Chaetomium globosum species complex. Persoonia. 2015; 36: 83–133.

12. Gonianakis M, Neonakis I, Darivianaki E, Gonianakis I, Bouros D, Kontou-Fili K. Airborne Ascomycotina on the island of Crete: seasonal patterns based on an 8-year volumetric survey. Aerobiologia 2005; 21: 69–74.

13. Samson RA, Houbraken J, Thrane U, Frisvad JC, Andersen B. Food and indoor fungi. In: CBS Laboratory Manual Series 2. CBS-KNAW Fungal Biodiversity Centre, Utrecht, The Netherland. 2010.

14. Sekita S, Yoshihira K, Natori S, Udagawa S, Muroi T, Sugiyama Y, et al. Mycotoxin production by Chaetomium spp. and related fungi. Canadian Journal of Microbiology 1981; 27: 766–772.

15. Seth HK. A monograph of the genus Chaetomium. Beihefte zur Nova Hedwigia 1970; 37: 1–133.

16. Andersen B, Frisvad JC, Søndergaard IIbS. Rasmussen I S. Larsen L S. Associations between fungal species and water-damaged building materials. Applied and Environmental Microbiology 2011; 77: 4180–4188.

17. Salo J.M, Kedves O, Mikkola R, Kredics L., Andersson, M.A, Kurnitski, J.; et al. Detection of Chaetomium globosum, Ch. cochliodes and Ch. rectangulare during the diversity tracking of mycotoxin-producing Chaetomium-like isolates obtained in buildings in Finland. Toxins 2020, 12, 443.

18. Wang X.W, Houbraken J, Groenewald JZ, Meijer M, Andersen B, Nielsen KF, Crous PW, Samson RA. Diversity and taxonomy of Chaetomium and Chaetomium-like fungi from indoor environments. Stud. Mycol. 2016, 84, 145–224.

19. Stiller MJ, Rosenthal S Summerbell RC., Pollack J., Chan A. Onychomycosis of the toenails caused by Chaetomium globosum. J. Am. Acad. Dermatol. 1992, 26, 775–776.

20. Aspiroz C, Gené J, Rezusta A, Charlez L., Summerbell RC. First Spanish case of onychomycosis caused by Chaetomium globosum. Med. Mycol. 2007, 45, 279–282.

21. Hwang S.M, Suh MK, Ha GY. Onychomycosis due to nondermatophytic molds. Ann. Dermatol. 2012, 24, 175–180.

22. Kim DM, Lee MH, Suh MK., Ha GY, Kim H, Choi JS. Onychomycosis caused by Chaetomium globosum. Ann. Dermatol. 2013, 25, 232–236.

23. Shi D, Lu G, Mei H, de Hoog G.S., Zheng H, Liang G., et al. Onychomycosis due to Chaetomium globosum with yellowish black discoloration and periungual inflammation. Med. Mycol. Case Rep. 2016, 13, 12–16

24. Hubka V, Mencl K., Skorepova M, Lyskova P, Zalabska E. Phaeohyphomycosis and onychomycosis due to Chaetomium spp., including the first report of Chaetomium brasiliense infection. Med. Mycol. 2011, 49, 724–733.

25. Lagace J, Cellier E. A case report of a mixed Chaetomium globosum/ Trichophyton mentagrophytes onychomycosis. Medical Mycology Case Reports 2012 ; 1: 76–78.

26. Latha R, Sasikala R, Muruganandam N, Shiva Prakash MR. Onychomycosis due to ascomycete Chaetomium globosum: a case report. Indian Journal of Pathology and Microbiology 2010; 53: 566–567.

27. Abbott SP, Sigler L, McAleer R, McGough DA, Rinaldi MG, Mizell G. Fatal cerebral mycoses caused by the ascomycete Chaetomium strumarium. J. Clin. Microbiol. 1995, 33, 2692–2698.

28. Thomas C, Mileusnic D., Carey RB, Kampert M, Anderson D. Fatal Chaetomium cerebritis in a bone marrow transplant patient. Hum. Pathol. 1999, 874–879.

29. Barron MA, Sutton DA, Veve R, Guarro J., Rinaldi M, Thompson E, et al. Invasive mycotic infections caused by Chaetomium perlucidum, a new agent of cerebral phaeohyphomycosis. J. Clin. Microbiol. 2003, 41, 5302–5307.

30. Mhmoud NA, Santona A, Fiamma M, Siddig EE, Deligios M, Bakhiet SM, et al. Chaetomium atrobrunneum causing human eumycetoma: The first report. PLoS Negl Trop Dis 2019; 13(5): e0007276. https://doi.org/10.1371/journal.

31. Dreyfuss M.. Taxonomische Untersuchungen inner-halb der Gattung Chaetomium. Sydowia 1976; 28:50–133.

32. Winter G. Pilze Ascomyceten. In: Rabenh Krypt Fl, 2nd ed (Leipzig) 1884–1887; 1(2):I–VI, 1–928.

33. Lee SJ, Hanlin RT. Phylogenetic relationships of Chaetomium and similar genera based on ribosomal DNA sequences. Mycologia 1999; 91:434–442, doi:10.2307/3761344.

34. Untereiner WA, De bois V, Naveau FA. Molecular systematics of the ascomycete genus Farrowia (Chaetomiaceae). Can J Bot 2001; 79:321–333, doi:10.1139/cjb-79-3-321

35. Zhang N, Castlebury LA, Miller AN, Huhndorf SM, Schoch CL, Seifert KA, et al. An overview of the systematics of the Sordariomycetes based on four-gene phylogeny. Mycologia 2006; 98: 1076–1087, doi:10.3852/mycologia.98.6.1076

36. Wang X, Zheng RY. Chaetomium acropullum sp. nov. (Chaetomiaceae, Ascomycota), a new psychrotolerent, mesophilic species from China. Nova Hedwigia 2005;80:413–7.

37. de Hoog GS, Ahmed SA, Najafzadeh MJ, Sutton DA, Keisari MS, Fahal AH, et al. Phylogenetic findings suggest possible new habitat and routes of infection of human eumyctoma. PLoS Neglected Tropical Diseases 2013 ; 7: e2229. http://dx.doi.org/10.1371/journal.pntd.0002229.

38. Davet P, Rouxel F. Detection and isolation of soil fungi. Enfield (NH): Science Publishers; 2000.

39. Warcup JH, Baker K.F. Occurrence of dormant ascospores in soil. Nature 1963; 197, 1317e1318.

40. Gunde-Cimerman N, Zalar P, Hoog GS de, Plemenitaš A. Hypersaline water in salterns – natural ecological niches for halophilic black yeasts. FEMS Microbiology Ecology 2000; 32: 235–240. Haubold EM, Aronson JF, Cowan DF, McGinnis

41. Möller EM, Bahnweg G, Sandermann H, Geiger HH. A simple and efficient protocol for isolation of high molecular weight DNA from filamentous fungi, fruit bodies, and infected plant tissues. Nucleic Acids Res 1992 ; 20:6115–6116

42. Nylander JAA. MrModeltest v. 2. Programme distributed by the author. Evolutionary Biology Centre, Uppsala University. 2004.

43. Burgess ND., Hales JD, Underwood E, Dinerstein E. Terrestrial Ecoregions of Africa and Madagascar: A Conservation Assessment; Island Press: Washington, DC, USA, 2004.

44. Pasternak Z., Al-Ashhab A, Gatica J, Gafny R, Avraham S, Minz D., et al. Spatial and temporal biogeography of soil microbial communities in arid and semiarid regions. PLoS ONE 2013, 8, e69705.

45. Silva M, Minter DW. Fungi from Brazil recorded by Batista and co-workers. Mycological Papers 1995; 169: 1–585.

46. Flavia R. Barbosa, Huzefa A. Raja, Carol A. Shearer, Luis F. P. Gusmão. Three Chaetomium species (Chaetomiaceae, Ascomycota) from the semiarid region of Brazil. Sitientibus série Ciências Biológicas 2012; 12(1): 115–118.

47. Kulneff C A. Comparative Study of Urban and Rural Diary Management Systems in Sudan. Uppsala. 2006.

48. Brausch G. Change and Continuity in the Gezira Region of the Sudan. Int Social Science J 1964; 16:340–356.

49. Carter A, Khan RS. New and interesting Chaetomium species from East Africa. Can J Bot 1982; 60:1253–1262.

50. Koch HA, Haneke H. Chaetomium funicolum Cooke als moglicher Erreger einer tiefen Mykose. Mkosen 1965; 9: 23–28.

51. Guppy KH, Thomas C, Thomas K, Anderson D. Cerebral fungal infections in the immunocompromised host: a literature review and a new pathogen Chaetomium atrobrunneum: case report. Neurosurgery 1998; 43:1463–1469.

52. Márquez SS, Bills GF, Zabalgogeazcoa I. The endophytic mycobiota of the grass Dactylis glomerata Fungal Diversity 2007; 27: 171–195.

53. Piepenbring M, Mendez O.A.C, Espinoza A.A.E, Kirschner R, Schofer H. Chromoblastomycosis caused by Chaetomium funicola: a case report from Western Panama. British Journal of Dermatology 2007; 157: 1025–1029.

54. Swarup G, Hansing ED, Rogerson CT. Fungi associated with sorghum seed in Kansas. Transactions of the Kansas Academy of Science 1962; 65(2): 120–137.

55. Matsushima T. Microfungi of the Solomon Islands and Papua-New Guinea. Publish by the author, Kobe. 1971.

56. Shearer CA, Crane JL. Illinois fungi XII. Fungi and Myxomycetes from wood and leaves submerged in southern Illinois swamps. Mycotaxon 1986; 25: 527–538.

57. Rai JN, Garg K, Jaitly AK.. Saprophytic fungi isolated from woods in mangrove swamps and their wood-decaying capability. Transactions of the Mycological Society of Japan 1981; 22(1): 65–74.

58. Cannon PF. A revision of Achaetomium, Achaetomiella and Subramaniula, and some similar species of Chaetomium. Trans Br Mycol Soc 1986; 87:45–76.

59. Rij JN, Wadhwani K., Tewari JP. Achaetomium macrosporum spec.nov. with notes on the genus Achaetomium. Indian Phytopathology 23, 54–57. Ranga Rao, V. & Mukerji, K. G. (1971a). Cytology of the ascus in Achaetomium globosum and A. luteum. Journal of General and Applied Microbiology 1970; 97, 311–318.

60. Rai JN,, Chowdhery HJ. Achaetomium uniapiculatum sp.nov., a new species of the genus Achaetomium. Current Science 1971; 40, 412–413..

61. Rai JN., Chowdhery HJ. Cytology of the ascus and ascocarp development in Achaetomium uniapiculatum. Journal of General and Applied Microbiology 1973; 19, 481–49.

62. Rai JN, Chowdhery HJ. Studies in the genus Achaetomium : two new species, A. sphaerocarpus and A. macrocarpus. Kavaka t, 1974a; 29–36.

63. Rai JN, Chowdhery HJ. Achaetomium fusisporus spec.nov, and A. sulphureus spec.nov. : two new species of the genus Achaetomium from Indian ‘Usar’ (alkaline) soils. Journal of the Indian Botanical Society S2, 1974b; 309–312.

64. Rai J.N. Chowdhery HJ. Achaetomium indicum Rai et Chowdhery spec.nov.: a new species of the genus Achaetomium from Indian ‘usar’ soils. Current Science 1978; 47, 23–24.

65. Rai JN, Saxena A. Cytology of the ascus and ascocarp development in Achaetomiella megaspore (Sorgel) D. Hawksw. Caryologia 1979; 32, 45–52

66. Locquin-Linard M. Achaetomium cristalliferum Faurel et Locquin-Linard. Nouvelle espece d’Ascornycete (Achaetomiaceae) isolee d’un sol aride. Cryptogamie, Mycologie 1980; 1, 235–240.

67. Udagawa S. A taxonomic study on the Japanese species of Chaetomium. Journal of General and Applied Microbiology 1960; 6, 223–251.

68. Udagawa S. A new species of Achaetomium. Transactions of the Mycological Society of Japan 1982; 23, 287–291.

69. Uoagawa, S, Sugiyama, Y. New records and new species of ascomycetous fungi from Nepal, a preliminary report on the expedition of 1980. Reports on the Cryptogamic Study in Nepal, March 1982 (Miscellaneous Publication of the National S cience Museum, Tokyo) 1982; 11–46.

70. Ara I, Sultana R, Chanda IF, Alam N. First report on Ovatospora brasiliensis from freshwater dried shrimp and prawn in Bangladesh. Int J Fauna Biol Stud 2020; 7: 43–47.

